# Left Behind: Modelling the life expectancy disparities amongst people with disabilities in Low and Middle-Income Countries

**DOI:** 10.1101/2023.07.12.23292565

**Authors:** Sara Rotenberg, Tracey Smythe, Hannah Kuper

**Affiliations:** University of Oxford, Oxford, OX1 2JD, United Kingdom; London School of Hygiene & Tropical Medicine, London, WC1E 7HT, United Kingdom; Stellenbosch University, Cape Town, 8000, South Africa

**Author notes:** corresponding author: Sara Rotenberg, University of Oxford, Oxford, OX1 2JD, United Kingdom. **Competing interests:** The authors declare no competing interests.

**Keywords:** disability, life expectancy, health equity

## Abstract

**Objective:** To use life tables to model the differences in life expectancy for people with and without disabilities in low- and middle-income countries (LMICs).

**Methods:** We used data from a recent conducted a meta-analysis of analysis which gathered data from 70 studies to determine Hazard Ratios (HRs) for all-cause mortality by disability status for children 0-15, adults 15-59, and adults 60+, using the World Health Organisation’s definition of disability. To assess the disparities in life expectancy among people with disabilities across 136 low and middle-income countries (LMICs), we constructed life tables using these HRs.gap compared to the population average. These calculations were based on the 2020 United Nations population projections. The life expectancy gap was meta-analysed across countries to calculate the mean difference.

**Findings:** People with disabilities in LMIC had a mean life expectancy of 49.3 years (95%C.I. 47.4 – 51.2), compared to 68.5 years (95% C.I. 67.4 – 69.5) for the general population, giving a median gap of 19.2 years (95% C.I. 18.3 – 20.1). The disparity in life expectancy varied across countries, ranging from 9.6 years (95% C.I. 4.7 – 17.4) in Bosnia and Herzegovina to 30.6 years (95% C.I. 16.9 – 40.6) in Nigeria.

**Conclusion:** Life expectancy inequities represent an urgent threat to upholding the rights of persons with disabilities and achieving global goals. It is crucial to address the disparities in social determinants of health, and prioritise the inclusion of people with disabilities within health equity efforts to close this gap.

## Background

Life expectancy is regarded as a core marker of health status, and comparisons between different groups have become a proxy for health equity.(1) Life expectancy has been increasing, with more than 20 years gained since the 1950s. However, these demographic gains are distributed unequally between countries and various social and marginalized groups.(2) Efforts to achieve health equity have used life expectancy to examine inequities based on race and geography, but there have been limited efforts to quantify these gaps by disability status, despite well-established health inequities for people with disabilities.(3-7) There are 1.3 billion people with disabilities globally (16% of the world’s population), making it important to understand potential inequities.(8) Moreover, 80% of people with disabilities live in low- and middle-income countries (LMICs), where inequities may be further exacerbated by inadequate access to healthcare and poverty.

People with disabilities are disproportionately affected by poverty,(9) exclusion from education and employment,(10) and social marginalisation,(11) which can contribute to poorer health status and outcomes, as well as lower life expectancy. In 2014, a UK Confidential Inquiry found that men and women with intellectual disabilities died 13 and 20 years earlier than the general population, respectively.(12) Several population-based studies and systematic reviews have echoed these findings for specific impairments. For instance, people with Down syndrome have 28 years shorter life expectancies than people without disabilities.(13) For psychosocial disabilities, several systematic reviews have put the life expectancy gap at 20.3 to 27.7 years for Schizophrenia globally (14) to 28.4 for any severe mental illness in Ethiopia. (15) These discrepancies are also observed with respect to physical impairments, as people with spinal cord injuries have 6.9 to 8.2 years shorter life expectancies, depending on the locality of their injuries.(16)

In addition to these impairment-specific studies, a few studies to date have also examined general functional difficulties and life expectancy in specific countries. For example Zheng et al, in China used a large-scale population-based dataset to show that men and women with any functional difficulties have 17.1 to 12.7 years shorter life expectancies, respectively.(17) Similarly, Bahk et al. also documented these differences in life expectancy, but also that they may be decreasing—from 20.4 year gap in 2004 to 16.4 year gap in 2017.(18) These findings are particularly alarming given the already low life expectancy in many LMICs. However, the needs and concerns of people with disabilities are often overlooked in national and international health programmes and investments, despite the significant health challenges that they face in these contexts.(8)

Nevertheless, there is limited global evidence on the life expectancies of people with disabilities in LMICs. We therefore sought to utilize new data on the higher mortality rates among people with disabilities(19) to model the life expectancy differences between people with disabilities and the general population in LMICs.

## Methods

### Definitions

For this analysis, we used the World Health Organization’s definition of disability as “an umbrella term for impairments, activity limitations, and participation restrictions”. (20) We considered life expectancy at birth, which is defined as the average number of years that a person can expect to live based on current age-specific mortality rates. Finally, we used the World Bank’s classification of countries based on their gross national income per capita to determine the sample of LMICs included in the model. As of 2021, the World Bank defined low-income countries as having a gross national income of $1,045 or less per capita and middle-income countries as having a gross national income between $1,046 and $12,735 per capita.(21)

### Data Sources

We obtained the 2020 UN population projections for LMIC from the United Nations Population Division.(22) Population and mortality data in one- and five-year cohorts were downloaded to create life tables. The data for children under five and adults over 90 were rearranged into the appropriate cohort categories (0, 1-4, and 90+) so they could match the cohorts used for analysis, while those aged five to 89 were copied verbatim. GDP per capita data was sourced from the World Bank country lending groups database.(21)

### Analysis

A systematic review and meta-analysis was conducted to estimate the mortality rate ratios of people with disabilities compared to those without by age group.(19) The systematic review included 70 studies conducted in 22 countries between 1990 and November 2022. These results were meta-analysed using a random effects model to calculate the Hazard Ratio (HR) for all-cause mortality by disability status. The pooled HR was estimated to be 4.46 (95% C.I. 3.01 – 6.59) for children 0-15, 3.53 (95% C.I. 1.29 – 9.66) for adults 15-59, and 1.97 (95% C.I. 1.63 – 2.38) for adults over 60.(19) We multiplied each cohort’s number of deaths from the life table by these HR estimates to estimate the number of deaths for people with disabilities, as well as a confidence interval for the number of deaths. This process was repeated for each country to create an estimate of life expectancy and a 95% confidence interval. Where the estimate for disabled deaths exceeded the population for that cohort, the number of deaths was replaced with the cohort’s total population, though this was only the case in five countries’ 90+ cohort for the upper confidence interval estimate, and so did not substantially impact our results.

Once this stage was completed, we constructed life tables for each country using the ‘phe_life_expectancy’ R package,(23) which uses the variance of life expectancy at each age group to calculate life expectancy at birth, as described by Chiang.(24) We produced two life tables. The first was for the general population (including people with and without disabilities), using the projected number of deaths, and the second was for the disabled population, which used the projected deaths multiplied by the HR for the respective age cohort. We then extracted the life expectancy at birth for the general population and the life expectancy at birth for disabled people to calculate the gap between the two for each country. Data were meta-analysed for all countries to calculate the mean and confidence intervals for life expectancy and the life expectancy gap. Associations were explored by country GDP per capita. R (Version 4.3.2) was used to conduct data analysis and create the figures.

## Results

Overall, people with disabilities had a median life expectancy of 49.3 years (95% C.I. 47.4 – 51.2), compared to 68.5 years (95% C.I. 67.4 – 69.5) for the general population. The mean life expectancy gap between people with disabilities and the general population was 19.2 years (95% C.I. 18.3 – 20.1).

The life expectancy gap was the highest in Nigeria (30.6 years, 95% C.I. 17.2 – 40.9), Chad (30.3 years, 95% C.I. 16.9 – 40.6), Somalia (30.1 years, 95% C.I. 16.7 – 41.5), South Sudan (30.0 years, 95% C.I. 16.3 – 41.3), and Guinea (29.1 years, 95% C.I. 15.7 – 41.8) and lowest in Bosnia and Herzegovina (9.6 years, 95% C.I. 4.7 – 17.5), Maldives (9.7 years, 95% C.I. 4.8 – 18), and North Macedonia (10.0 years, 95% C.I. 4.7 – 18.6). Figure 1 shows a map summarizing the results from the life expectancy gap between people with disabilities and the general population across 136 LMICs (Supplementary Table 1).

**Fig. 1:**
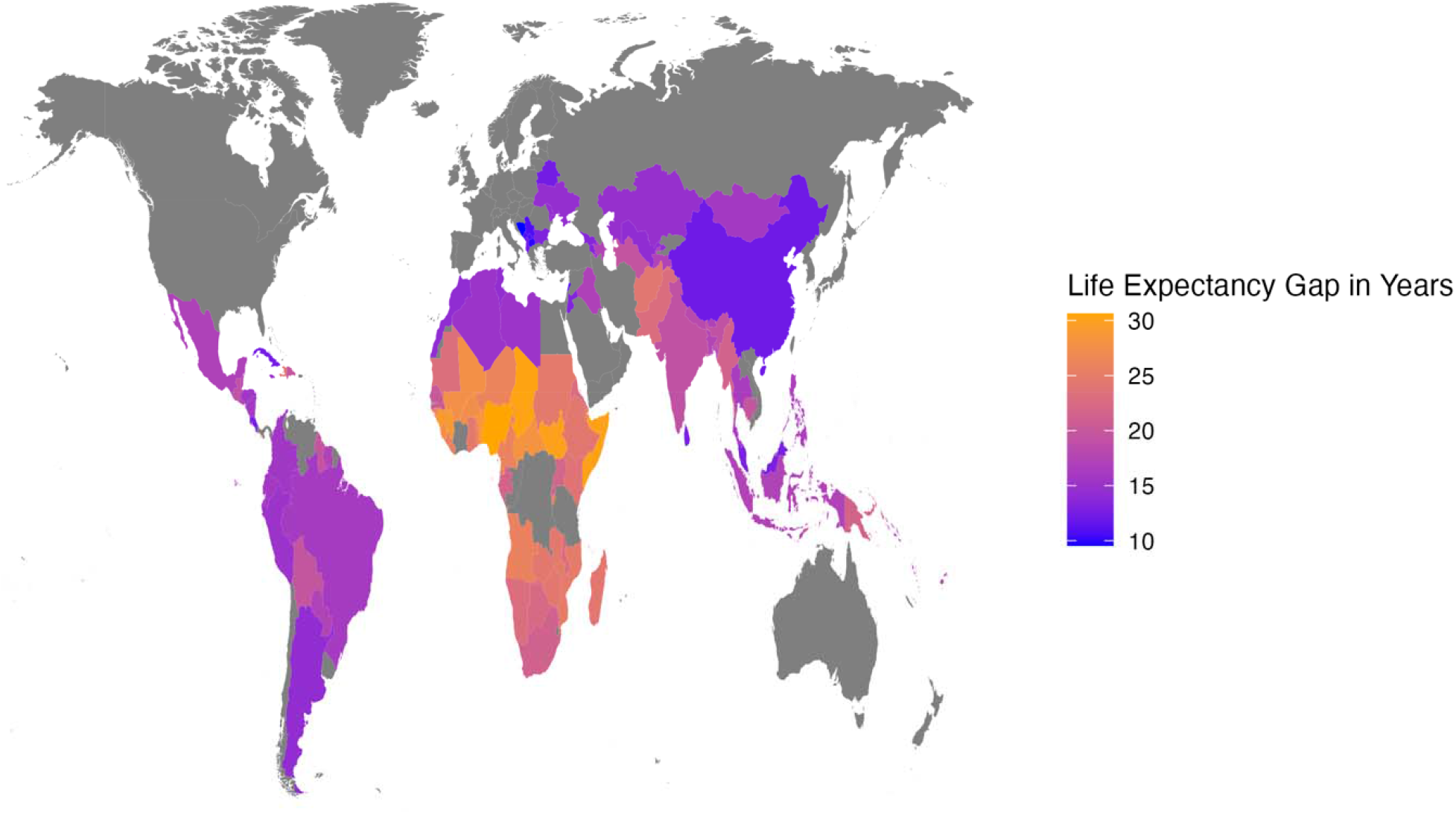
Life Expectancy Gap in 136 low- and middle-income countries

**Fig. 2:**
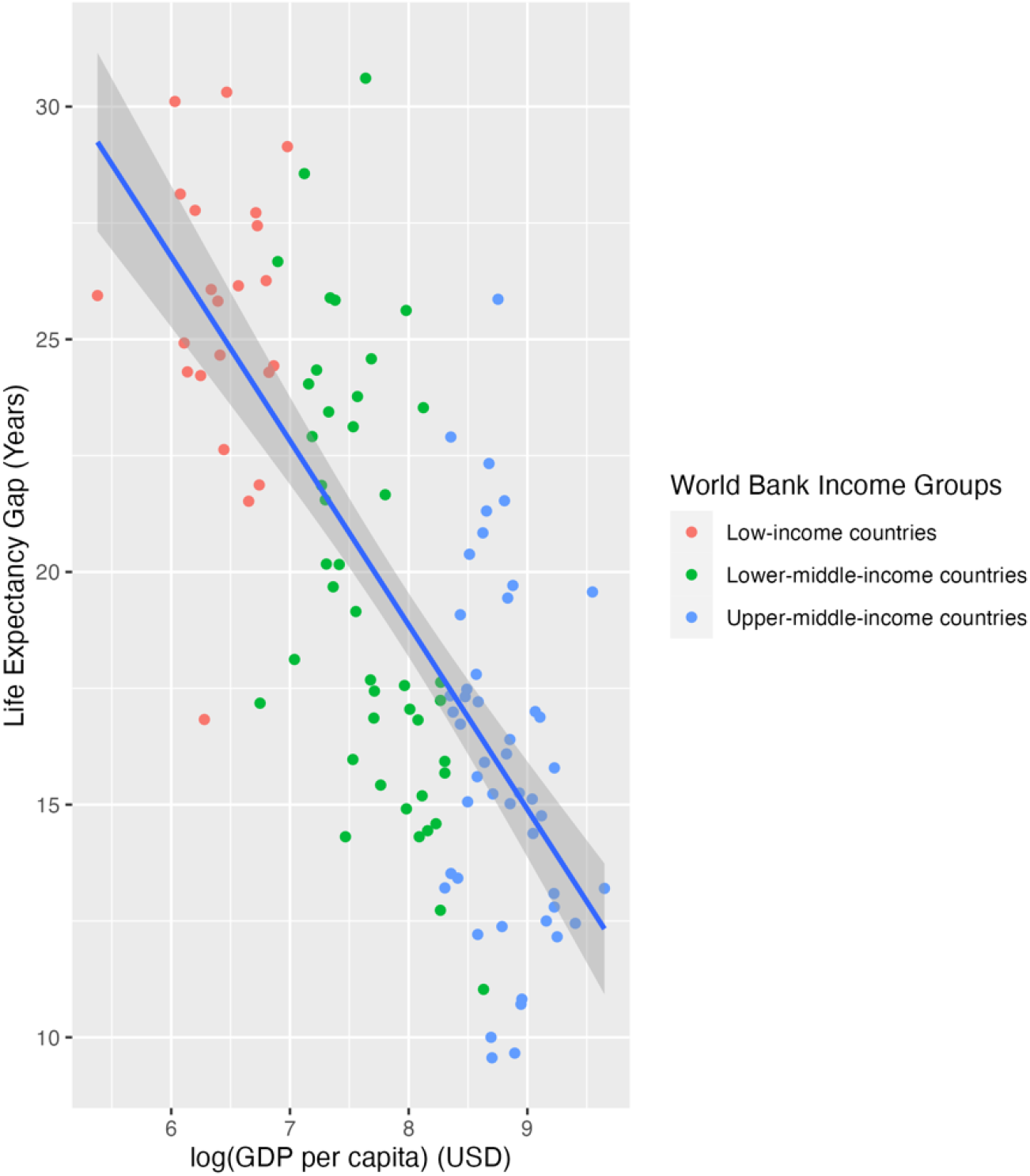
Life Expectancy Gap by GDP per capita (USD)

Table 1 shows the differences in life expectancy by country income grouping. In these aggregate measures, there is a higher gap in low-income countries (26.1 years, 95% C.I. 13.0-39.8) and the lowest gap in lower-middle upper-middle-income countries (14.6 years, 95% C.I. 6.3 – 27.3).

**Table 1:** Life expectancy by disability status and life expectancy gap for people with disabilities by World Bank Income Group

|  | General Population Life Expectancy | Life Expectancy for People with Disabilities [95% C.I.] | Life Expectancy Gap [95% C.I.] |
| --- | --- | --- | --- |
| Low-income countries | 62.9 | 36.8 [23.1 – 49.9] | 26.1 [13.0 – 39.8] |
| Lower-middle income countries | 68.2 | 46.3 [32.5 – 57.5] | 21.9 [10.7 – 35.7] |
| Upper-middle income countries | 75.8 | 61.3 [48.5 – 69.5] | 14.6 [6.3 – 27.3] |
| All LMICs* | 68.5 | 49.3 [47.4 – 51.2] | 19.2 [18.3 – 20.1] |

\* calculated using inverse variance meta-analysis

There was an overall association between GDP and the life expectancy gap, and this is particularly strong for lower-middle income countries. However, some countries diverted from this trend, including Nigeria (30.6 years, 95% C.I. 17.2 – 40.9), a lower-middle income country with the highest life expectancy gap, and Benin, which had a life expectancy gap of 28.6 years (95% C.I. 15.1 – 41.5). The life expectancy gap by GDP per capita is demonstrated in Figure

## Discussion

This paper provides novel insight into the potentially substantial gaps in life expectancies for people with disabilities. Our model uses life tables, population projections, and hazard ratios from a recent systematic review to show that people with disabilities have a mean life expectancy gap of 19.2 years (95% C.I. 18.3 -20.1) compared to the general population across all LMICs. Moreover, our findings suggest that the life expectancy gaps for people with disabilities is associated with a country’s income status, as there exists a negative correlation between GDP per capita and the life expectancy gap in this population.

Our models provide simplistic and illustrative rather than definitive estimates. Nevertheless, our results are consistent with data from the around the world show that people with disabilities experience a life expectancy gap of 10-20 years, regardless of geography or impairment type (Table 2) (5) (12-18, 25, 26). While the majority of population-based studies are from high-income countries, our model-generated estimates demonstrate notable similarities in available LMIC data. Specifically, our modelled estimates from China (12.2 years, 95% C.I. 5.5-23.2 years) and Ethiopia (24.3, 95% C.I. 11.6 – 38.7) are within the range reported by Zheng et al. (12.7 for females and 17.1 for males with physical disability)(17) and Fekadu et al. (28 year gap for people with severe mental illness) (15), respectively. It is noteworthy that our model does not disaggregate by impairment type due to data limitations, although the studies also suggest there are particularly high gaps in life expectancy for people with mental illness and intellectual disabilities (12-15, 25), as demonstrated in Table 2.

**Table 2:**
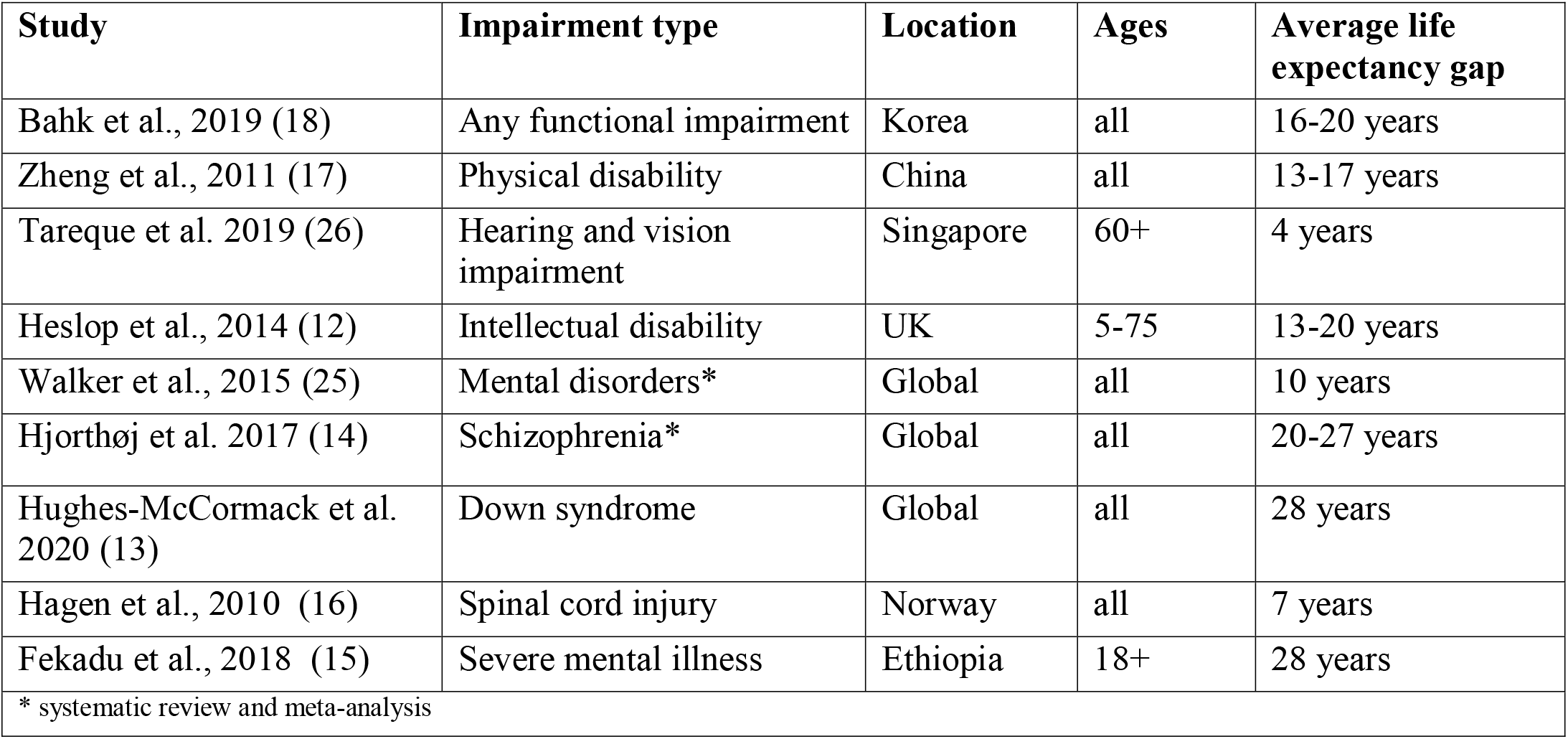
Life Expectancy Gap Papers

| Study | Impairment type | Location | Ages | Average life expectancy gap |
| --- | --- | --- | --- | --- |
| Bahk et al., 2019 (18) | Any functional impairment | Korea | all | 16-20 years |
| Zheng et al., 2011 (17) | Physical disability | China | all | 13-17 years |
| Tareque et al. 2019 (26) | Hearing and vision impairment | Singapore | 60+ | 4 years |
| Heslop et al., 2014 (12) | Intellectual disability | UK | 5-75 | 13-20 years |
| Walker et al., 2015 (25) | Mental disorders* | Global | all | 10 years |
| Hjorthøj et al. 2017 (14) | Schizophrenia* | Global | all | 20-27 years |
| Hughes-McCormack et al. 2020 (13) | Down syndrome | Global | all | 28 years |
| Hagen et al., 2010 (16) | Spinal cord injury | Norway | all | 7 years |
| Fekadu et al., 2018 (15) | Severe mental illness | Ethiopia | 18+ | 28 years |
| * systematic review and meta-analysis |  |  |  |  |

Whilst a component of the shorter life expectancy for people with disabilities can be attributed to the underlying health condition or impairment of people with disabilities, it is unlikely to be the sole reason. Indeed, the Confidential Inquiry in the UK showed that overall 37% of avoidable deaths among people with intellectual disabilities were from causes amenable to good quality healthcare. Various social determinants of health are therefore also likely factors that contribute to these disparities. For instance, people with disabilities may face socioeconomic disadvantage that can negative impact their health status and access to healthcare, such as lower incomes, lack of education and poor social support. (9) People with disabilities, including when seeking healthcare. These barriers can lead to inadequate care, mistrust of medical professionals, or lack of access to healthcare services.(27, 28) Consequently, discrimination and stigma is likely contribute to the overall health disparities and increase the risk of premature death for people with disabilities.

Yet, there still needs to be understanding of the causal relationship between poor access to healthcare and premature mortality for people with disabilities so that it can be mitigated. One way that poor access to healthcare can lead to premature mortality for people with disabilities is through delayed or missed diagnoses. People with disabilities often face barriers to accessing healthcare, such as lack of transportation, inaccessible medical facilities, and lack of healthcare providers trained to care for people with disabilities. (3-5) These barriers can lead to delayed or missed diagnoses of serious health conditions, which can increase the risk of premature death. Another issue is the lack of preventive care for people with disabilities. Regular preventive care, such as cancer screenings, vaccinations, and health screenings (6, 29, 30), may not be readily available to this population, increasing the risk of preventable illnesses and premature death.

Inadequate treatment for health conditions is another consequence of poor access to healthcare for people with disabilities. People with disabilities may not receive appropriate treatment for their health conditions or prevent them from seeking care in the future due to a lack of accessible healthcare services (31) or lack of healthcare providers trained to care for people with disabilities (32). This inadequate treatment can lead to the progression of serious health conditions, which can increase the risk of premature death. It is important to recognize that poor access to healthcare can interact with and exacerbate other factors that contribute to premature mortality, such as poverty and lack of education, making the problem even more complex.

Policymakers and stakeholders must prioritise the development of disability-inclusive health policies and programs to address these gaps in life expectancy. It is crucial to acknowledge that accepting significantly shorter lives for people with disabilities is unacceptable. Efforts should focus on addressing the underlying determinants of health and implementing disability-inclusive strategies. This requires a collaborative, multi-sectoral approach involving the health, social, education, and employment sectors, among others, to foster disability inclusion and enhance the health and well-being of people with disabilities on a global scale. Change is needed across the health system, including improving fundamental building blocks such as governance, leadership, and financing, as well as service-delivery components of healthcare worker skills on disability, accessibility of facilities, and availability of treatments.(5) Intervening in these components to promote disability-inclusion offers opportunities to improve health access for this population and close the gap.(5) These changes are also likely to be generate high returns on investment, and improve the health system across the population.(8)

### Strengths and Limitations

Our approach enables the estimation of differences in life expectancy for people with disabilities compared to the general population by country using readily available data sources, using best available data from a recent systematic review and meta-analysis. It is the first study to examine life expectancy gaps for people with disabilities in more than one country and model the life expectancy gap for this population.

However, it is important to acknowledge that models provide simplistic estimations and that more robust data is needed on life expectancy by disability status. There are a number of limitations to consider. First, the models take account of the demographic makeup of each country, but it cannot fully account for various other social factors that impact life expectancy, such as race, gender, or rurality. Second, not all countries included in this modelling exercise reported data for the systematic review, and, therefore, the estimated association of disability and mortality may not be accurate for these particular countries. Third, it is likely that the life expectancy gap was underestimate, as our model assessed the gap in life expectancy between people with disabilities and the general population, yet on average 16% of people with disabilities in the general population will have disabilities. Furthermore, many of the studies included in the review over-adjusted the associations measured (e.g. adjusted for poverty thus underestimating the “true” relationship of disability and mortality), which would also contribute towards our model underestimating the life expectancy gap. Future research should explore the complex interactions between disability, health, and social determinants, and develop interventions that can effectively reduce the disability-mortality gap in LMICs.

## Conclusion

Our results reinforce the urgent need for policymakers and stakeholders to prioritize disability-inclusive health policies and programs that address these underlying determinants of health. Put simply, the status quo accepts that people with disabilities die much younger, on average. Without invested efforts for improvement, we will continue to view these gaps in life expectancy as expected, acceptable differences for people with disabilities. Instead, we should use these gaps to motivate investment into equitable health and social systems that support and uplift the rights of people with disabilities.

## Data Availability

All data produced in the present study are available upon reasonable request to the authors

